# Pan-cancer analyses of the associations between 109 pre-existing conditions and cancer treatment patterns across 19 adult cancers

**DOI:** 10.1101/2022.11.11.22282219

**Authors:** Wai Hoong Chang, Alvina G. Lai

## Abstract

Comorbidities present considerable challenges to cancer treatment and care. However, little is known about the effect of comorbidity on cancer treatment decisions across a wide range of cancer types and treatment modalities. Harnessing a cohort of 280,543 patients spanning 19 site-specific cancers, we explored pan-cancer frequencies of 109 comorbidities. Multinomial regression revealed that patients with comorbidity exhibited lower odds of receiving chemotherapy and multimodality treatment. End-stage renal disease was significantly associated with a decreased odds of receiving chemotherapy and surgery. Patients with prostate cancer who have comorbid non-acute cystitis, obstructive and reflux uropathy, urolithiasis, or hypertension were less likely to receive chemotherapy. Among patients with breast cancer, dementia, left bundle branch block, peripheral arterial disease, epilepsy, Barrett’s oesophagus, ischaemic stroke, unstable angina and asthma were associated with lower odds of receiving multimodal chemotherapy, radiotherapy and surgery. Comorbidity is also consistently associated with the lower odds of receiving chemotherapy when comparing across 10 drug classes. Patients with comorbid dementia, intracerebral haemorrhage, subarachnoid haemorrhage, oesophageal varices, liver fibrosis sclerosis and cirrhosis and secondary pulmonary hypertension were less likely to receive antimetabolites. Comorbidity can influence the effectiveness and tolerability of cancer treatment and ultimately, prognosis. Multi-specialty collaborative care is essential for the management of comorbidity during cancer treatment, including prophylactic measures to manage toxicities.

## Introduction

Patients with cancer do not usually have cancer alone. Comorbidities present challenges to cancer treatment because they influence treatment decisions, prognosis and the overall trajectory of care. Yet, the influence of specific comorbidities at various points of the cancer care pathway is not well understood. Certain comorbidities might not directly influence survival but could affect patients’ tolerability to treatment. Cancer therapy could also exacerbate pre-existing conditions or give rise to new comorbidities due to treatment-related toxicities. In the UK, the National Institute for Health and Care Excellence (NICE) guidelines for cancer management have limited information on comorbidities. The guideline for lung cancer briefly mentioned that radiotherapy with curative intent should consider the presence of comorbidities[1]. The NICE breast cancer guideline stated that patients with invasive breast cancer should be treated with surgery and systemic therapy, irrespective of age unless significant comorbidity precludes surgery[2]. There is no mention of what significant comorbidity entails. The guideline also recommends offering trastuzumab to patients with invasive breast cancer and to consider comorbidities but did not specify which comorbidities to consider nor did it mention how comorbidities should be managed during cancer treatment. Similarly, the prostate cancer guidelines stated that docetaxel chemotherapy should only be offered to patients who do not have significant comorbidities but did not mention specific comorbidities[3]. Guidelines emphasise the importance of considering comorbidities in treatment planning, yet provide little information on how comorbidities should be handled.

Randomised controlled trials (RCTs) are essential for the investigation of the safety and efficacy of new cancer drugs while offering patients the opportunity to access experimental therapeutics. The number of RCTs has steadily increased over the years, however, inclusion and exclusion criteria have mostly remained unchanged. RCTs often exclude large segments of cancer patients with comorbidities[4]. The presence of one or more comorbidities is associated with a decreased odds of trial participation and trial offers[5]. An inevitable consequence is that RCTs create study cohorts that are divergent from real-world populations and produce limited data on the tolerability of therapies in people with comorbidities.

Due to limitations in clinical guidelines and trial evidence, relatively little is known about the influence of comorbidity on the choice of cancer treatment. Harnessing real-world cancer populations, we aim to address this gap by characterising real-world treatment patterns in patients with comorbid conditions. We performed a population cohort study using linked electronic health records (EHRs) from primary care, secondary care and the cancer registry. We explored 7 treatment categories and 10 chemotherapy drug classes across 19 adult cancers. Specific objectives are (i) to estimate the frequencies of 109 comorbidities diagnosed within the 5-year period before cancer diagnosis across 19 cancer types, (ii) to examine patterns of multimorbidity across cancer types, (iii) to estimate the associations between comorbidity and cancer treatment decisions and (iv) to estimate the associations between comorbidity and chemotherapy use across 10 chemotherapy drug classes. As cancer survivorship increases, the generation of a new knowledge base demonstrating the impact of comorbidity on cancer treatment could inform multidisciplinary team meetings to improve overall safety and optimise care.

## Methods

### Data sources

We employed linked electronic health records from primary care, secondary care and the cancer registry in England. Primary care data were recorded using Read and SNOMED codes. Secondary care Hospital Episode Statistics data were recorded in ICD-10. Detailed information on cancer, tumour stage, grade and tumour count were obtained from the National Cancer Registration and Analysis Service (NCRAS). Treatment information and chemotherapy drug details were obtained from the NCRAS systemic anti-cancer treatment and radiotherapy datasets. Socioeconomic deprivation statuses were obtained from the Office for National Statistics. Socioeconomic deprivation was recorded using Index of Multiple Deprivation which is a measure of relative deprivation based on 37 indicators and seven domains to identify the most and least deprived areas[6]. Information governance approval was received from the Medicines and Healthcare products Regulatory Agency (19222).

### Study design and electronic health record phenotypes

Patients with incident site-specific cancer aged 18 years or older were identified during the study period of 01-01-1998 to 31-10-2020. We considered 19 site-specific cancers: (i) bladder, (ii) brain, (iii) breast, (iv) cervix, (v) colon and rectum, (vi) gallbladder and biliary tract, (vii) kidney, (viii) liver and intrahepatic bile duct, (ix) lung, (x) melanoma, (xi) oesophagus, (xii) oropharynx, (xiii) ovary, (xiv) pancreas, (xv) prostate, (xvi) stomach, (xvii) testis, (xviii) thyroid and (xix) uterus. Only patients with cancer treatment information were included. We considered 7 treatment categories: (i) surgery alone, (ii) chemotherapy alone, (iii) radiotherapy alone, (iv) chemotherapy and surgery, (v) radiotherapy and surgery, (vi) chemotherapy and radiotherapy and (vii) chemotherapy, radiotherapy and surgery. We analysed ten chemotherapy drug categories: (i) alkylating agents, (ii) anthracyclines, (iii) antimetabolites, (iv) biological response modifiers including immunotherapy, (v) hormonal agents, (vi) kinase inhibitors, (vii) non-anthracycline antitumour antibiotics, (viii) plant alkaloids excluding vinca alkaloids, (ix) platinum agents and (x) vinca alkaloids.

Phenotypes were obtained from an open-access library[7] and have been previously validated[8]. We considered 109 non-cancer comorbidities categorised into nine organ systems: cardiovascular (33 conditions), endocrine (5 conditions), gastrointestinal (18 conditions), haematological (8 conditions), immunology and infection (19 conditions), musculoskeletal (8 conditions), neurological (10 conditions), pulmonary (4 conditions) and renal (4 conditions). We considered comorbidities that were diagnosed in the 5-year period before cancer diagnosis to rule out historical diagnoses that may have been cured.

### Statistical analyses

The primary outcomes were cancer treatment and chemotherapy drug type, given a specific comorbidity. Treatment outcomes were identified from the NCRAS dataset. The proportions of patients with specific comorbidities were estimated by cancer type. The number of comorbidities (0, 1, 2, 3, 4, 5 and 6+) was also summarised by cancer type. As the dependent variable (treatment type) had 7 categories, multinomial logistic regression was employed to ascertain the association between comorbidity and cancer treatment, adjusting for age, sex, socioeconomic status, tumour grade, tumour stage, tumour count (according to the NCRAS data dictionary, this is the count of every tumour for a patient) and multimorbidity count. Comorbidity (independent variable) was defined as the presence or absence of a particular condition (i.e., with heart failure vs. no heart failure). Odds ratios (ORs) and 95% confidence intervals (CIs) for the odds of receiving a particular treatment given a comorbidity were calculated. Multinomial logistic regression results for treatment decisions were presented relative to surgery alone as the baseline choice of treatment.

For the association between comorbid conditions and chemotherapy type, as the dependent variable (chemotherapy type) had two categories (e.g., received or did not receive a particular chemotherapy drug), binomial logistic regression models were fitted. Binomial logistic regression models were adjusted for cancer type, age, sex, socioeconomic status, tumour grade, tumour stage, tumour count and multimorbidity count. For binomial regression, comorbidity was used as the independent variable and was defined as the presence or absence of a particular condition.

Variance inflation factors (VIFs) were calculated to test for multicollinearity of the independent variables in regression models. Variables with VIF greater than five were removed from the model. All analyses were performed according to the STROBE guidelines. All analyses were performed in accordance with the relevant guidelines and regulations. All analyses were performed using R 3.6.3 with the following packages: tidyverse, data.table, tableone, nnet, questionr and performance.

## Results

We identified 280,543 patients having an incident cancer diagnosis with cancer treatment information (Table S1). Of these, 131,528 (46.9%) were male. Patients within each age group were as follows: age 18-34 (5,127; 1.8%), age 35-50 (29,553; 10.5%), age 51-65 (89,511, 31.9%), age 66-80 (120,080; 42.8%) and age ≥ 81 (36,272; 12.9%). Patients were categorised into seven treatment groups: surgery alone (117,007; 41.7%), chemotherapy alone (30,007; 10.7%), radiotherapy alone (36,100; 12.9%), chemotherapy and radiotherapy (18,326; 6.5%), chemotherapy and surgery (24,396; 8.7%), radiotherapy and surgery (35,121; 12.5%) and chemotherapy, radiotherapy and surgery (19,586; 7.0%). 57,384 patients had information on the types of chemotherapy that were prescribed (Table S2). The following chemotherapy types were considered: alkylating agents (8,224; 14.3%), anthracyclines (8,960; 15.6%), antimetabolites (21,224; 37.0%), biological response modifiers including immunotherapy (7,638; 13.3%), hormonal agents (30,795; 53.7%), kinase inhibitors (1,785; 3.1%), non-anthracycline antitumour antibiotics (791; 1.4%), plant alkaloids excluding vinca alkaloids (12,819; 22.3%), platinum agents (18,490; 32.2%) and vinca alkaloids (2,147; 3.7%). The proportions of patients receiving each of the seven forms of therapy and each of the ten chemotherapy drug types for across cancer types are shown in Table S3 and Table S4, respectively.

### Patterns of recently diagnosed conditions across 19 adult cancers

We analysed the proportions of 109 conditions (grouped into 9 organ systems) that were diagnosed in the 5-year period before cancer diagnosis. The full list of conditions is provided in Table S5. We observed a high variability in the diagnosis of comorbid conditions across cancer types. Among patients with brain cancer, the top three comorbidities were intracranial hypertension (14.03% [95% CI: 13.01-15.05]), hypertension (13.65% [12.64-14.66]) and epilepsy (10.96% [10.04-11.88]) (Figure 1, Table S6). The top three comorbidities for colorectal cancer were peritonitis (15.25% [14.92-15.59]), hypertension (13.51% [13.19-13.83]) and anaemia (12.31% [12.01-12.62]). For gallbladder and biliary tract cancer, the top comorbidities were cholangitis (49.96% [47.20-52.72]), cholelithiasis (30.95% [28.39-33.50]) and cholecystitis (30.71% [28.16-33.26]). For liver and intrahepatic bile duct cancer, the top comorbidities were hepatic failure (20.12% [18.26-21.99]), fatty liver (19.51% [17.66-21.35]), cholangitis (19.28% [17.45-21.11]) and portal hypertension (16.64% [14.91-18.37]). Common comorbidities in patients with lung cancer include lower respiratory tract infections (18.75% [18.31-19.18]), chronic obstructive pulmonary disease (15.78% [15.37-16.18]) and hypertension (15.28% [14.88-15.68]). For oesophageal cancer, the top comorbidities were oesophagitis and oesophageal ulcer (18.14% [17.30-18.97]), hypertension (13.31% [12.57-14.04]) and Barrett’s oesophagus (12.92% [12.19-13.64]). The full results for all cancers are available in Table S6 and graphically represented in Figure 1 and Figure S1.

**Figure 1.**
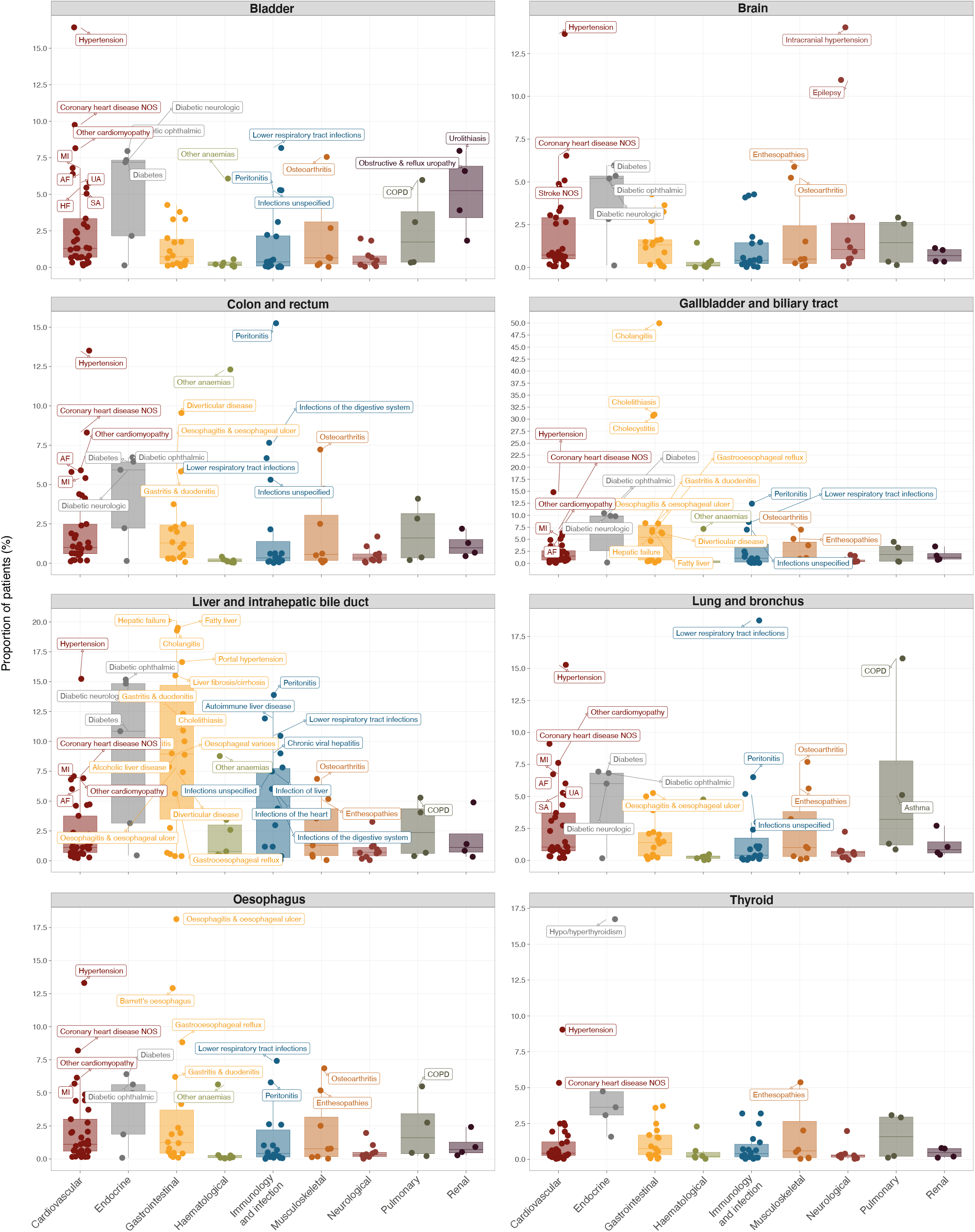
Non-cancer conditions diagnosed before cancer. Boxplots show the proportion of patients having a first diagnosis of any of the 109 non-cancer conditions (grouped into 9 organ systems) in the 5-year period before cancer diagnosis. Boxplots for 8 cancer types are shown in this figure. Conditions with proportions ≥ 5% are annotated on the plots. Boxplots for the remaining 11 cancer types are shown in Figure S1. Full data and confidence intervals are presented in Table S6.

### Multimorbidity burden in patients with cancer

Pan-cancer analyses of the number of comorbidities revealed that patients with certain cancers experienced a high proportion of pre-existing conditions. Patients with liver, gallbladder, pancreatic or lung cancers had the highest number of comorbidities when considering 109 conditions (Figure 2, Table S7). The proportion of patients with six or more co-morbid conditions were as follows: liver cancer (63.07% [60.83-65.31]), gallbladder cancer (53.22% [50.46-55.98]), pancreatic cancer (47.46% [46.03-48.90]) and lung cancer (41.69% [41.14-42.24]) (Table S7). By contrast, 45.43% [43.54-47.32] of patients with testicular cancer had no comorbid conditions. A reasonable number of patients with cervical cancer (27.66% [26.17-29.15]) or melanoma (20.96% [20.34-21.59]) also had zero comorbidities. See Table S7 for the full results.

**Figure 2.**
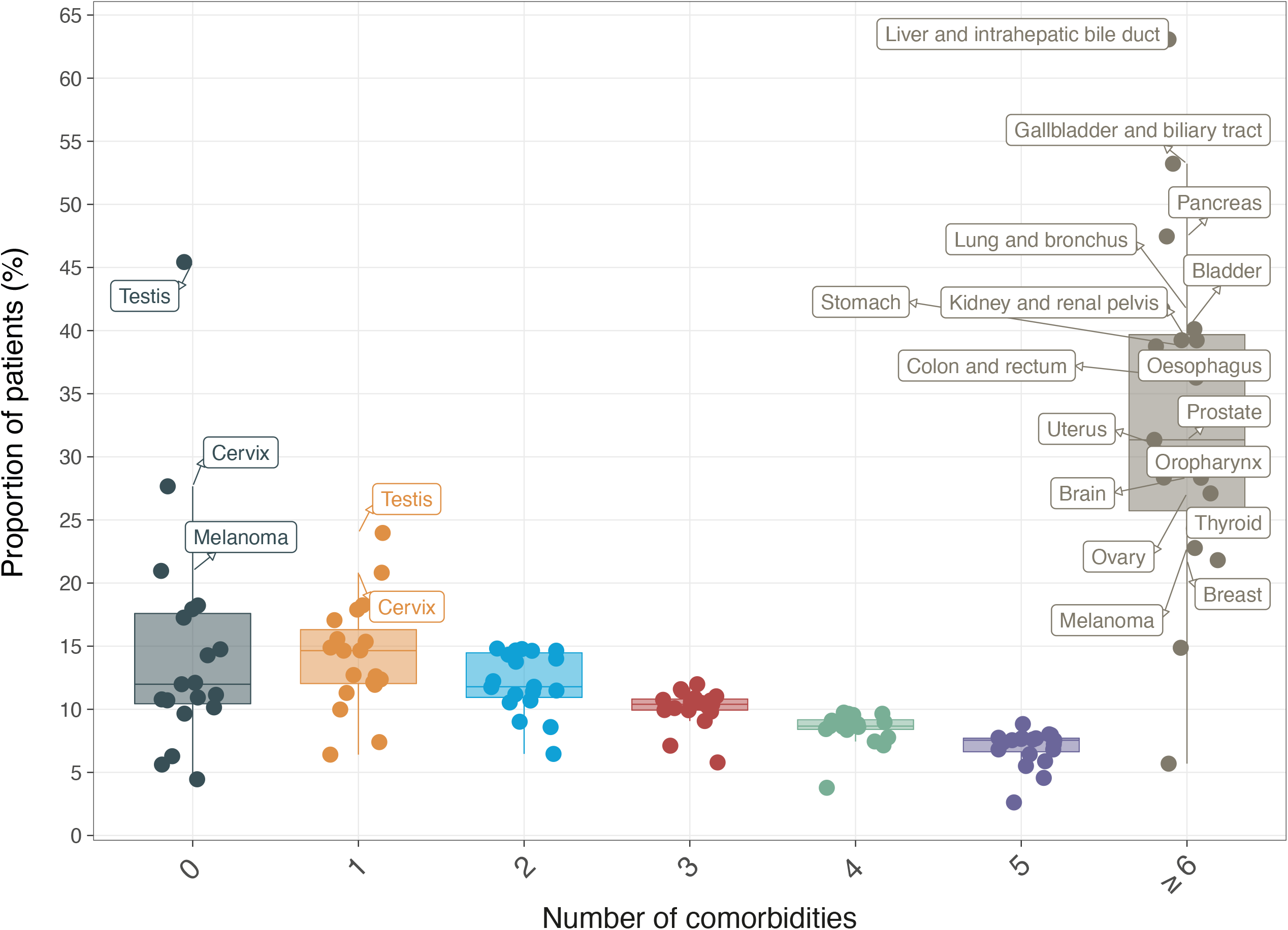
Multimorbidity patterns across 19 adult cancers. Boxplot depicts the proportion of patients, within each cancer type, having 0, 1, 2, 3, 4, 5 or ≥ comorbidities. Cancers with proportions ≥ 20% are annotated on the plot. Full data and confidence intervals are presented in Table S7.

### Associations between comorbidity and cancer treatment decisions

We examined the relationship between comorbid conditions and the type of cancer treatment using multinomial logistic regression, with ‘surgery alone’ as the baseline choice of treatment. Adjusted odds ratios (ORs) for the odds of receiving a particular treatment combination given a specific comorbidity were calculated (Table S8). ORs with P value < 0.05 are graphically represented in Figure 3 and Figures S2 to S9; all results are presented in Table S8. Patients with cardiovascular conditions had lower odds of receiving (i) chemotherapy alone, (ii) chemotherapy and radiotherapy, (iii) chemotherapy, radiotherapy and surgery, (iv) chemotherapy and surgery and (v) radiotherapy and surgery (Figure 3, Table S8). For example, patients with colorectal cancer having comorbid atrial fibrillation were less likely to receive chemotherapy, radiotherapy and surgery (OR 0.55 [CI: 0.40-0.75]), chemotherapy and radiotherapy (0.63 [0.47-0.83]), chemotherapy and surgery (0.69 [0.58-0.81]), chemotherapy alone (0.69 [0.57-0.82]) and radiotherapy and surgery (0.80 [0.66-0.96]) (Figure 3, Table S8).Patients with heart failure were also less likely to receive chemotherapy and radiotherapy; ORs for breast (0.47 [0.32-0.68]), oesophageal (0.48 [0.27-0.85]), lung (0.63 [0.50-0.79]), colorectal (0.63 [0.46-0.87]) and prostate (0.74 [0.57-0.95]) cancers. Similarly, patients with colorectal (0.42 [0.28-0.64]), breast (0.51 [0.39-0.67]) and prostate (0.57 [0.40-0.79]) cancers who also have heart failure were less likely to receive chemotherapy, radiotherapy and surgery.

**Figure 3.**
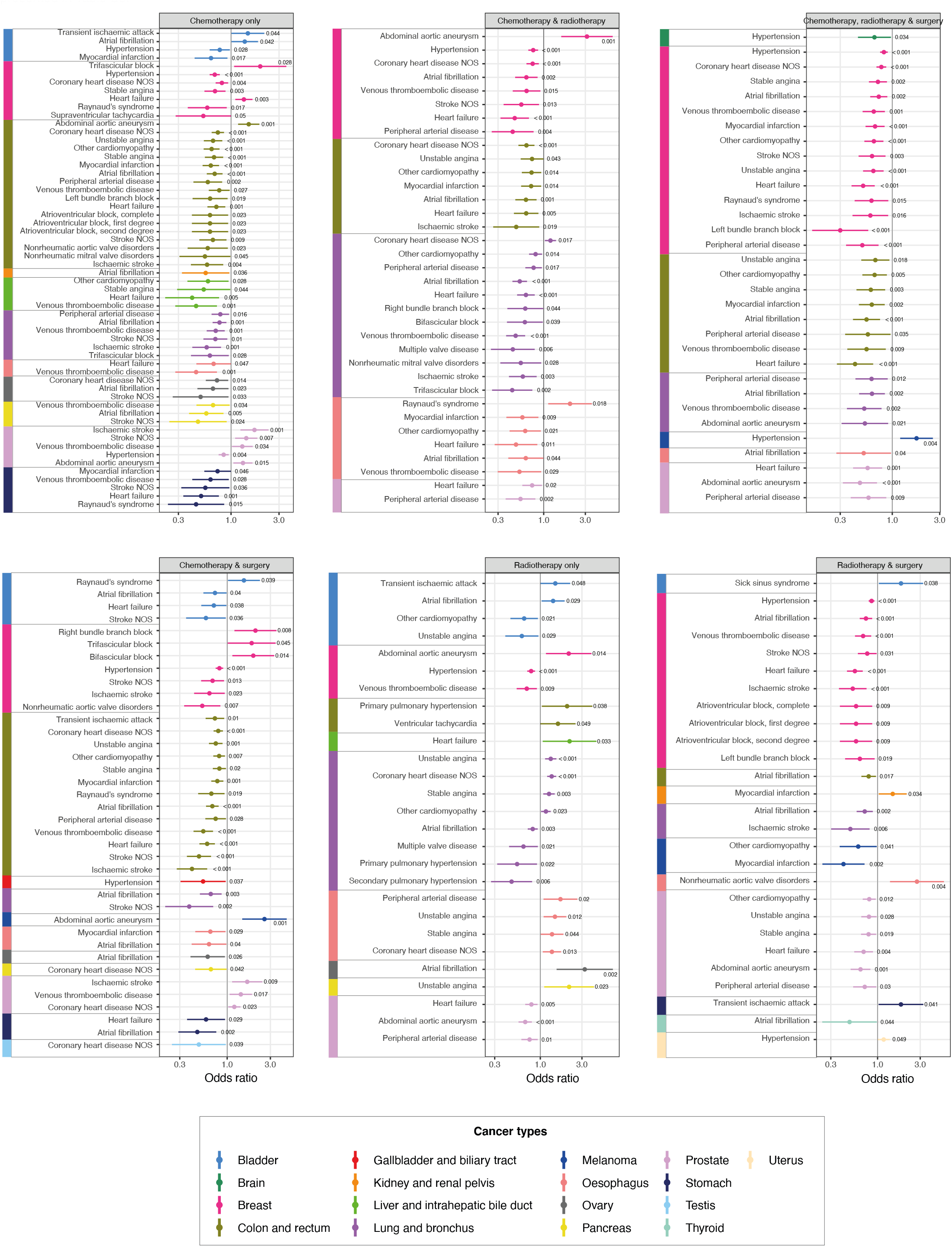
Multinomial logistic regression investigating the associations between cardiovascular conditions and cancer treatment decisions across adult cancers. Forest plots show odds ratios for a particular treatment type, adjusted for age, sex, socioeconomic status, tumour grade, tumour stage, tumour count and multimorbidity count. Seven treatment categories were considered (i) surgery alone, (ii) chemotherapy alone, (iii) radiotherapy alone, (iv) chemotherapy and radiotherapy, (v) chemo-therapy and surgery, (vi) radiotherapy and surgery, and (vii) chemotherapy, radiotherapy and surgery. Multinomial logistic regression models were fitted using surgery alone as the baseline choice of treatment for each cancer type (colour-coded). Only cardiovascular conditions are shown in this figure. Plots for the remaining 8 organ systems are shown in Figures S2 to S9. Only results with P < 0.05 are shown in the figure. P values are annotated on the plots. Full data and confidence intervals are presented in Table S8.

When considering gastrointestinal conditions, patients with cholelithiasis were less likely to receive chemotherapy alone (relative to surgery alone); ORs for gallbladder (0.44 [0.30-0.67]), liver (0.50 [0.35-0.72]) and prostate (0.61 [0.44-0.85]) cancers (Figure S3, Table S8). Additionally, patients with kidney (0.44 [0.25-0.78]), colorectal (0.62 [0.52-0.74]), lung (0.83 [0.71-0.97]) or prostate (0.86 [0.74-0.99]) cancers with comorbid diverticular disease of the intestine were less likely to receive radiotherapy alone. When considering renal comorbidities, end-stage renal disease was significantly associated with a decreased odds of receiving chemotherapy and surgery; ORs for oesophageal (0.36 [0.19-0.69]), colorectal (0.48 [0.37-0.64]), lung (0.50 [0.32-0.77]) and bladder (0.62 [0.42-0.89]) cancers (Figure S9, Table S8). Similarly, patients with lung (0.43 [0.26-0.70]), colorectal (0.47 [0.28-0.80]), prostate (0.50 [0.34-0.74]) or breast (0.70 [0.51-0.97]) cancers who have comorbid end-stage renal disease were less likely to receive all three forms of treatment (i.e., chemotherapy, radiotherapy and surgery).

When exploring specific cancer types, patients with prostate cancer who have comorbid non-acute cystitis (OR 0.30 [0.16-0.54]), obstructive and reflux uropathy (OR 0.45 [0.34-0.59]), urolithiasis (OR 0.48, [0.38-0.60]) or hypertension (OR 0.84 [0.75-0.95]) were less likely to receive chemotherapy alone, relative to surgery alone (Table S8). For patients with breast cancer, dementia (OR 0.25 [0.15-0.44]), left bundle branch block (OR 0.30 [0.16-0.57]), peripheral arterial disease (OR 0.50 [0.34-0.73]), epilepsy (OR 0.51 [0.29-0.92]), Barrett’s oesophagus (OR 0.58 [0.40-0.85]), ischaemic stroke (OR 0.60 [0.40-0.91]), unstable angina (OR 0.65 [0.51-0.82]) and asthma (OR 0.74 [0.63-0.87]) were associated with lower odds of receiving multimodal chemotherapy, radiotherapy and surgery (Table S8).

### Associations between comorbidity and chemotherapy decisions

We next investigated the odds of receiving a particular chemotherapy drug given a specific comorbidity using binomial logistic regression. Adjusted ORs for all comorbidities are shown in Table S9. ORs with P value < 0.05 are graphically represented in Figure 4 and Figure S10; all results are presented in Table S9. Cancer patients with comorbidities compared with those without comorbidities were less likely to receive antimetabolites (Figure 4, Table S9). The top 10 conditions were dementia (OR 0.19 [0.14-0.25]), intracerebral haemorrhage (0.31 [0.22-0.42]), subarachnoid haemorrhage (0.31 [0.21-0.43]), oesophageal varices (0.35 [0.22-0.52]), liver fibrosis sclerosis and cirrhosis (0.40 [0.31-0.51]), secondary pulmonary hypertension (0.41 [0.26-0.61]), primary pulmonary hypertension (0.41 [0.27-0.61]), ischaemic stroke (0.42 [0.35-0.51]), other interstitial pulmonary diseases with fibrosis (0.51 [0.37-0.67]) and venous thromboembolic disease (0.51 [0.46-0.57]). Similarly, patients with comorbidities were less likely to receive platinum-based chemotherapy: dementia (0.15 [0.11-0.21]), primary pulmonary hypertension (0.31 [0.19-0.47]), secondary pulmonary hypertension (0.31 [0.19-0.48]), intracerebral haemorrhage (0.38 [0.28-0.50]), oesophageal varices (0.38 [0.23-0.58]), subarachnoid haemorrhage (0.41 [0.30-0.55]), liver fibrosis sclerosis and cirrhosis (0.43 [0.33-0.55]), ischaemic stroke (0.47 [0.39-0.56]), glomerulonephritis (0.48 [0.37-0.63]) and scoliosis (0.51 [0.33-0.75]). Patients with comorbidities were also less likely to receive kinase-targeted therapy: ischaemic stroke (0.42 [0.22-0.70]), stroke (not otherwise specified) (0.45 [0.27-0.71]), obstructive and reflux uropathy (0.48 [0.25-0.81]), lower respiratory tract infections (0.53 [0.45-0.63]), peripheral arterial disease (0.54 [0.35-0.79]), venous thromboembolic disease (0.55 [0.40-0.73]), Raynaud’s syndrome (0.56 [0.35-0.85]), end stage renal disease (0.61 [0.45-0.80]), heart failure (0.63 [0.47-0.81]) and myocardial infarction (0.63 [0.49-0.81]) (Figure S10, Table S9).

**Figure 4.**
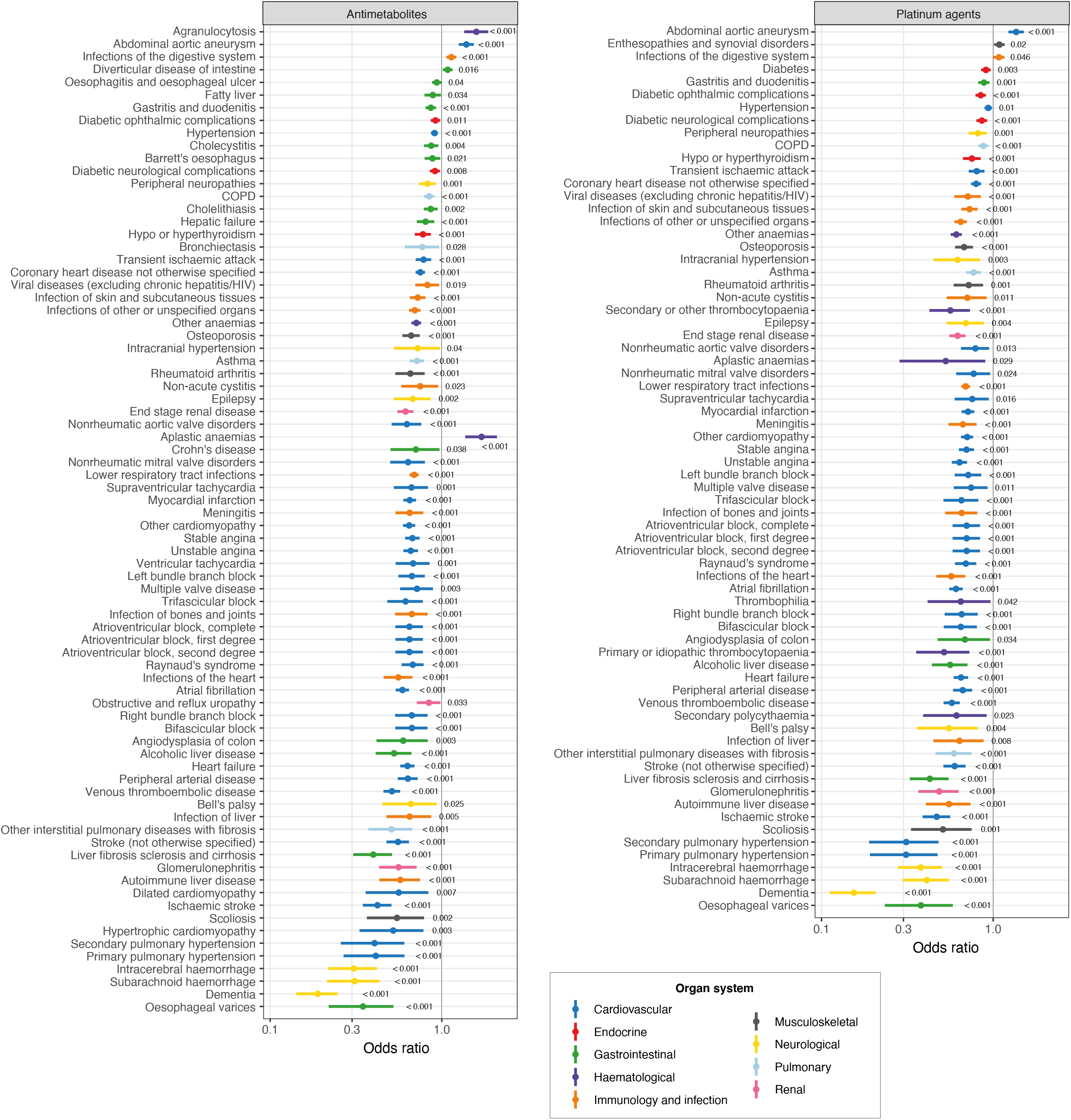
Binomial logistic regression investigating the associations between comorbidity and chemotherapy decisions. Forest plots show odds ratios for a particular chemotherapy type, adjusted for cancer type, age, sex, socioeconomic status, tumour grade, tumour stage, tumour count and multimorbidity count. Ten chemotherapy classes were considered – results for antimetabolites and platinum agents are shown in this figure. Plots for the remaining 8 chemotherapy classes are shown in Figure S10. Binomial logistic regression models were fitted. Conditions were colour-coded according to the 9 organ systems. Only results with P < 0.05 are shown in the figure. P values are annotated on the plots. Full data and confidence intervals are presented in Table S9.

## Discussion

This pan-cancer population-based study involving 19 adult cancers fills an evidence gap by investigating the associations between a wide range of comorbid conditions and cancer treatment patterns. For the first time, we investigated 109 comorbidities involving 9 organ systems and found that the type and magnitude of comorbidity burden were variable across cancers. For example, patients with liver cancer had a high burden of comorbidity involving a wide range of cardiovascular, infection, endocrine and gastrointestinal conditions. Given that the risk factors of gastrointestinal cancers include obesity, infection, alcohol abuse and smoking, a high burden of comorbidity was similarly observed in other gastrointestinal cancers (i.e., pancreatic, stomach, colorectal, gallbladder and oesophageal cancers). Pre-existing conditions were associated with a reduced likelihood of receiving multimodality therapy. Patients with comorbidities were less likely to receive chemotherapy; this pattern is consistent across the 10 chemotherapeutic agents we considered.

Comorbid conditions affect treatment choices in several ways. Our observations on the negative association between comorbid conditions and receipt of chemotherapy are consistent with other studies. A systematic review of 16 studies found 11 studies reporting that patients with comorbidities were less likely to be given chemotherapy[9]. Ten studies estimated the odds of chemotherapy use and except for a single study, they all reported decreased use of chemotherapy with ORs ranging from 0.25 to 0.99, regardless of cancer stage or tumour site[9]. Furthermore, a study on ovarian cancer demonstrated longer delays in initiating chemotherapy in patients with comorbidity[10]. A Canadian study on 20,689 patients with lung cancer found that pre-existing cardiovascular disease was associated with a lower likelihood of receiving chemotherapy (OR 0.53 [0.48-0.58])[11]. A Dutch study on stage III colon cancer reported that patients with comorbidity received chemotherapy less often (OR 0.6 [0.5–0.8])[12]. Functional limitations and geriatric syndromes were associated with a lower likelihood of receiving chemotherapy and surgery, but only if patients have two or more comorbidities[13]. Advanced age, presence of comorbidities and poor performance status were common reasons for withholding adjuvant chemotherapy in patients with stage III colorectal cancer[14]. Patients who were given chemotherapy were more likely to develop complications (52%) than those receiving surgery alone (41%), suggesting high toxicity rates due to interacting comorbidities[14]. Women with breast cancer and severe comorbidity were less likely to receive both chemotherapy and radiotherapy[15]. Women with severe comorbidity were also less likely to be offered the hormonal therapy tamoxifen (OR 0.88 [0.78-0.99])[15]. Breast-conserving surgery is less likely to be given to women with a high comorbidity burden (OR 0.63 [0.58-0.69])[15]. In patients with colorectal cancer, comorbidities were associated with a greater likelihood of receiving surgery alone[13]. Patients with stage III colon cancer with a Charlson Comorbidity Index score of ≥ 3 were less likely to be given chemotherapy[16]. Comorbidity, in patients with ovarian cancer, was associated with a lower likelihood of receiving standard combination chemotherapy (OR 0.03 [0.01-0.1])[17]. Patients with multimorbidity and those with functional limitations were 40% less likely to receive chemotherapy[18]. Age is a strong predictor of comorbidity and could, in part, explain the influence on cancer treatment choices. Elderly patients with colon cancer were less likely to receive antimetabolite chemotherapy (5-fluorouracil)[19]. Elderly patients with lung or head and neck cancer were also less likely to receive standard treatment[20, 21]. Elderly patients with comorbidities were less likely to adhere to chemotherapy due to potential toxicities[22]. We found that the associations between comorbid conditions and treatment decisions persisted even after adjusting for age, and other relevant prognostic factors such as tumour stage, grade, tumour count, sex and socioeconomic status.

Previous studies investigating the impact of comorbidity on treatment choice are heterogeneous in design and methodology, limited to specific cancer types and treatment modalities. Index systems such as the Charlson Comorbidity Index (CCI) have been developed to generate an overall comorbidity score based on a list of conditions. CCI was created to predict the risk of death within 1 year of hospitalisation. However, CCI also includes solid tumours, leukaemia and lymphoma in its list of conditions, which might not add additional information if applied to patients with cancer. A cancer-specific Comorbidity Index was subsequently developed using prostate and breast cancer cases from the Surveillance, Epidemiology, and End Results Program dataset[23]. This index system uses conditions in CCI but excludes cancer conditions. Adult Comorbidity Evaluation 27 (ACE-27) is a 27-item comorbidity index designed using hospital-based cancer registries, which does not take into account information from primary care[24]. Validations of ACE-27 were also undertaken using data from a single medical site[24]. We demonstrated that the pattern of comorbidities is variable across cancer types. This means that each comorbidity might exert varying prognostic impact and the use of general index systems may result in limited specificity. We have taken a disease agnostic view to consider all comorbidities diagnosed in the preceding five years before cancer diagnosis. Most studies are limited by the sources of data used in the development of general or disease-specific comorbidity measures. To overcome this limitation, we have harnessed population health records across England rather than using a single disease population.

### Challenges in considering the role of comorbidity in cancer treatment decisions

Decisions to offer a specific course of treatment and the suitability of the treatment for patients are often based on perceived benefits and risks. The presence of significant comorbidities or poor performance status are indicators for withholding therapy in patients with advanced cancer[25]. Over 50% of patients with advanced lung cancer receive no treatment, which is concerning. Among those receiving treatment, 94% of first-line therapy was platinum-based chemotherapy. Nonetheless, patients with poor performance status experienced a significant survival benefit when treatment is provided, which contradicts the notion that reduced functional capacity should be a criterion for deciding against treatment[25]. However, performance status, in this case, might not be a suitable prognostic measure for deciding treatment plans. Performance status is vulnerable to subjective evaluation, with demonstrable observer biases[26, 27]. The presence of specific comorbidities should be considered instead, and further evaluation of safety profiles should be conducted in specific patient populations.

Nephrotoxicity of chemotherapeutic agents has been well documented[28]. We observed that chemotherapy is less likely to be given to patients with end-stage renal disease. Cisplatin is primarily eliminated by the kidneys and it’s recommended that a lower dose should be used in patients with renal impairments[29]. Patients with end-stage renal disease receive dialysis, however, haemodialysis and plasmapheresis are ineffective in eliminating cisplatin to reverse overdose[30]. Lung resection provides the highest chance of cure in patients with lung cancer. However, patients with chronic obstructive pulmonary disease (COPD) and more severe airway obstruction were less likely to undergo thoracic surgery with curative intent (OR 0.025 [0.004-0.167])[31]. Additionally, the risks of postoperative pulmonary complications such as empyema and atelectasis are higher in patients with COPD[32, 33]. Comorbidity can also exacerbate treatment-related complications. Patients with diabetes face a greater risk of complications from breast cancer treatment, including neuropathy, heart disease, poor wound healing, increased susceptibility to infection and nephropathy[34]. Diabetes is associated with ipsilateral upper arm dysfunction five years after mastectomy[35]. Diabetes is also linked to an increased risk of complications from radiotherapy. Patients with cervical cancer and comorbid diabetes experience bowel obstruction and rectovaginal fistula[36]. Over 60% of patients with ovarian cancer and diabetes who received paclitaxel or cisplatin chemotherapy experience worsening hyperglycaemia and progression of neurological symptoms[37].

### Management of comorbidity during and after cancer treatment

Radiotherapy and certain chemotherapy agents can adversely affect the vascular system and heart. Patients with pre-existing cardiovascular disease or cardiovascular risk factors are at an even greater risk of developing cardiac complications. Radiation can affect the valves, vascular structures and pericardium or myocardium[38, 39]. Monitoring for cardiovascular function during anticancer therapy could reduce adverse effects. Radiation induces the fibrosis and calcification of valvular tissue. Valvular dysfunction induced by radiation has a median time to diagnosis of two decades after radiotherapy[40]. At the time of valvular disease diagnosis, most patients are no longer under the care of oncologists, thus the accuracy of detecting this condition becomes limited if cancer treatment is not captured in detail in the medical records[41]. For symptomatic patients, the American Society of Echocardiography and the European Association of Cardiovascular Imaging recommend yearly physical and clinical history examination of echocardiography[42]. For asymptomatic patients, transthoracic echocardiogram 10 years after radiotherapy is recommended.

Collaborative cardiac and cancer care is associated with improved outcomes and better cardioprotection. Consultations with cardiologists were associated with a higher frequency of heart failure medication prescription and better survival outcomes[43]. Chemotherapy-induced left ventricular systolic dysfunction is diagnosed, on average 2 years after cancer diagnosis, suggesting that regular echocardiography surveillance may help identify late-manifesting cardiac injury[43]. Baseline cardiovascular risk assessment (electrocardiogram, left ventricular ejection fraction [LVEF], cardiac biomarkers and lipid panel) before anticancer therapy could lessen the chances of developing cardiovascular complications[44]. Anthracyclines and certain targeted therapies (e.g., trastuzumab or sorafenib) are associated with a high risk of left ventricular dysfunction or heart failure. Patients with pre-existing cardiovascular disease who received trastuzumab, doxorubicin or both show sustained decline in LVEF over 3 years post-therapy[45]. Beta-blockers, angiotensin-converting enzyme inhibitors or angiotensin receptor blockers can reduce the risk of cardiotoxicity in these patients[46].

### Implications for practice

The role of comorbidity should be considered at different stages along the cancer care pathway. Due to the advancements in cancer treatment, more and more patients are surviving cancer and comorbidity may increase the risk of developing late effects[47, 48] during the survivorship period. Comorbidity may increase the risk of certain cancers and the risk of recurrence. Comorbid conditions may affect cancer diagnostic timeliness, participation in cancer screening programmes and consequently stage at diagnosis. Comorbidities exert varying effects on help-seeking behaviour. Patients with lung cancer who have COPD took longer to consult with symptoms of lung cancer[49]. Similarly, patients with dementia are less likely to seek help for cancer symptoms[50]. Comorbidity is associated with a longer diagnostic interval from the first presentation to cancer diagnosis and delayed diagnosis is often linked to a higher tumour stage at diagnosis[51]. Comorbidity influences treatment choice, adherence to treatment, surveillance post-treatment and overall prognosis. Comorbidity may influence the decision to offer adjuvant chemotherapy if there is a perceived risk for such treatment to affect the underlying condition. Comorbidity may increase treatment toxicity or might be a reason for withholding curative-intent or multimodality treatment, thus making it challenging to determine whether poor survival is due to the comorbidity or patients receiving less intensive therapy. Considering specific comorbidities individually rather than composite measures could be more useful to assess how they might affect cancer treatment, subsequent monitoring and overall outcome. It is also important to disentangle the impact of comorbidity on treatment decisions, i.e., clinicians not offering certain treatments for patients with certain comorbidities (physician factors) versus patients with a high degree of functional impairment refusing treatment due to previous health experience (patient factors). Age and other patient characteristics may also modify the relationship between comorbidity and cancer treatment decisions. To design an optimal treatment plan for patients, a data-informed systematic approach to assess the risks posed by individual comorbidities on specific treatment regimens is warranted. The definition of comorbidity is not consistent across studies, which does not allow for cross-comparisons and precludes meta-analysis. Some studies use disease-specific measures while others use index systems. Our study seeks to address this limitation by evaluating the impact of individual comorbidities, using a systematic and consistent methodology, across 19 cancer types.

### Strengths and limitations

To our knowledge, this is the first population cohort study investigating the impact of 109 conditions on cancer treatment decisions across 19 adult cancers, employing a consistent methodology allowing for cross-comparison. We employed linked data from primary care, hospitals and cancer registry. The use of routinely collected health records means that this study is not affected by biases inherent to self-reported measures. Previous studies on comorbidities often rely on general measures (e.g., CCI), which assumes that they have a similar impact in different disease populations. We have taken a different approach to estimate the effects of single conditions on cancer treatment choice to improve prognostic utility. Other factors can influence preferences for treatment and treatment outcomes. We account for heterogeneity among patients by considering age, sex, socioeconomic status, tumour grade, tumour stage, tumour count and the presence of multiple comorbid conditions in regression models.

We acknowledge several limitations. Although this is a retrospective cohort study, it provides a real-world depiction of treatment patterns in patients across diverse cancer types. We relied on diagnoses recorded in ICD-10, Read and SNOMED codes and have not quantified the severity of individual conditions. Nonetheless, despite this limitation, our study provides a comprehensive assessment of comorbidities, and the use of population-based cohorts helps ensure the generalisability of results. The data also reflects conditions that are managed in both community-based general practices and conditions requiring hospitalisation. We have not explored the impact of specific combinations of comorbidities on treatment decisions. We also did not explore the reciprocal effects of how cancer diagnosis or treatment could influence the management of comorbidities, which is beyond the scope of this study but may have implications on overall patient prognosis.

### Conclusion

Comorbidity affects cancer treatment decisions. It can interact with cancer and influence the tolerability or effectiveness of cancer treatment. A thorough assessment of comorbid conditions, especially renal, liver and cardiac function, in every patient prior to starting cancer treatment would allow for dosage adjustment of anticancer drugs. Where nephrotoxic or cardiotoxic anticancer therapy is required, preventive measures such as hyper-hydration, diuresis or managing modifiable cardiovascular risk factors through drug prophylaxis may be warranted[52, 53]. Patients with comorbidity require care by different physicians, which often occurs at different time points and involves different specialties and settings. To optimise care, effective information exchange and interprofessional or cross-sectoral collaboration are required. Patients may be offered a more active role in healthcare coordination. By keeping patients informed of their status, it could facilitate information flow between all providers of care[54].

## Data Availability

All data produced in the present work are contained in the manuscript.

